# Optimizing antiviral treatment for seasonal influenza in the United States: A Mathematical Modeling Analysis

**DOI:** 10.1101/2020.07.28.20163741

**Authors:** Matan Yechezkel, Martial L Ndeffo Mbah, Dan Yamin

**Affiliations:** Department of Industrial Engineering, Tel Aviv University, 55 Haim Levanon St, Tel Aviv, Israel; College of Veterinary Medicine & Biomedical Sciences, College Station, Texas, 77843; School of Public Health, Texas A&M University, College Station, Texas, 77843

**Keywords:** Seasonal Influenza, Antiviral treatment, Vaccination, Mathematical modeling

## Abstract

Seasonal influenza remains a major health burden in the United States. Despite recommendations of early antiviral treatment of high-risk patients, the effective treatment coverage remains very low. We developed an influenza transmission model that incorporates data on infectious viral load, social contact, and healthcare-seeking behavior, to evaluate the population-level impact of increasing antiviral treatment timeliness and coverage among high-risk patients in the US. We found that increasing the rate of early treatment among high-risk patients who received treatment more than 48 hours after symptoms onset, would substantially avert infections and influenza-induced hospitalizations. We found that treatment of the elderly has the highest impact on reducing hospitalizations, whereas treating high-risk individuals aged 5-19 years old has the highest impact on transmission. The population-level impact of increased timeliness and coverage of treatment among high-risk patients was observed regardless of seasonal influenza vaccination coverage and the severity of the influenza season.

## Introduction

Seasonal influenza continues to be a major cause of health and economic burden (Molinari et al., 2007). Although influenza infection is generally a self-limiting disease, it can result in severe illness and death. In particular, the disease carries substantial health burden among young children, the elderly, and people with certain health conditions (Fiore et al., 2010; Monto, 2008). In the United States, seasonal influenza results in an estimated incidence of 9.3–49.0 million illnesses, 140,000–710,000 hospitalizations, and 12,000–56,000 deaths annually (Molinari et al., 2007; Tokars et al., 2018a).

Vaccination is the mainstay of efforts to reduce the burden of seasonal influenza. The US Advisory Committee on Immunization Practices (ACIP) recommends influenza vaccination for all individuals aged six months or older. However, the majority of the US population does not comply with these recommendations, and vaccination rates against seasonal influenza hover around approximately 40% annually (Chiu et al., 2017). Additionally, due to the rapid mutation of the virus and an imperfect match between the vaccine’s virus cocktail, vaccine efficacy is not complete and varies widely by season. For example, the average influenza vaccine effectiveness was estimated to be 45% in the US, with the annual value ranging between 19% and 60% over the past decade (“CDC Seasonal Flu Vaccine Effectiveness Studies | CDC,” n.d.; Doyle et al., 2019).

Recently, increased severity of influenza, marked by high rates of outpatient and inpatients visits, has been observed among patients with high-risk influenza-associated complications (Garten et al., 2018; Xu et al., 2019). This group includes children under five years old, adults over 65 years old, American Indian/Alaska natives, pregnant women, people with immunosuppression, people who are morbidly obese, people with chronic pulmonary or cardiovascular conditions, and people with diabetes (Fiore et al., 2011). This high-risk population accounts for the bulk of influenza-associated hospitalizations in the US. For example, more than 50% of all influenza-associated hospitalizations in the US occur among adults over 65 years old, and more than 70% of adult inpatients have at least one underlying medical condition that placed them at high risk for influenza-associated complications (Appiah et al., 2015; Blanton et al., 2017; Davlin et al., 2016; Garten et al., 2018).

The increase in influenza-associated hospitalization and mortality (“Influenza-associated Pediatric Mortality,” n.d.; Xu et al., 2019) has raised concerns about vaccine uptake and early treatment of patients at risk of severe complications (Ison, 2018; O’Halloran et al., 2016). Neuraminidase inhibitors (NAIs) are a class of antiviral medications recommended for the pharmacologic treatment of influenza (Fiore et al., 2011). Early therapy of influenza patients with NAIs reduces the duration and intensity of viral shedding, the duration of symptoms, and disease-associated complications, hospitalizations, and mortality (Hayden and Pavia, 2006). Despite the high burden of influenza-induced complications among high-risk individuals, their rate of treatment for influenza infection has remained low (Stewart et al., 2018). Approximately 40% of high-risk patients with laboratory-confirmed influenza seek care within two days of symptoms onset (Stewart et al., 2018). Among these patients, on average, 37% are prescribed an antiviral medication (Biggerstaff et al., 2014; Stewart et al., 2018). The ACIP guidelines recommend that antiviral treatment be given to high-risk patients with clinically suspected influenza infection, even with deferred laboratory confirmation, when influenza is known to be circulating in the population (Fiore et al., 2011). However, current clinical practice is far from keeping with these guidelines.

Antiviral treatment not only provides direct benefits to treated patients by reducing their risk of influenza-induced hospitalization and/or mortality but may also provide indirect protection to noninfected individuals by reducing their risk of infection. This indirect benefit is achieved by decreasing the contribution of treated patients to disease transmission by reducing their viral shedding and duration of infectiousness. Household-based trials have shown that early treatment of infected individuals with NAIs may reduce their contribution to disease transmission by 50 – 80% (E. et al., 2009; Halloran et al., 2007).

To evaluate the population-level impact of increased antiviral treatment coverage and timeliness of influenza-infected high-risk individuals during influenza seasons, we developed a data-driven influenza transmission model that incorporates data on infectious viral load, social contact, healthcare-seeking behavior, time to seek healthcare, and antiviral treatment.

## Methods

### Model Overview

We developed a dynamic model for influenza infection progression and transmission in Texas, California, Connecticut, and Virginia. Our model is a modified susceptible-infected-recovered compartmental framework (Vynnycky and White, 2010), in which transitions between the health-related compartments occur over time (Figure 1). To model age-dependent transmission, we stratified the population into five age groups: 0–4 y, 5–19 y, 20–49 y, 50–64 y, and ≥ 65 y. We also distinguished between high-risk and low-risk individuals for each age group based on the ACIP case definition (Fiore et al., 2011).

**Figure 1.**
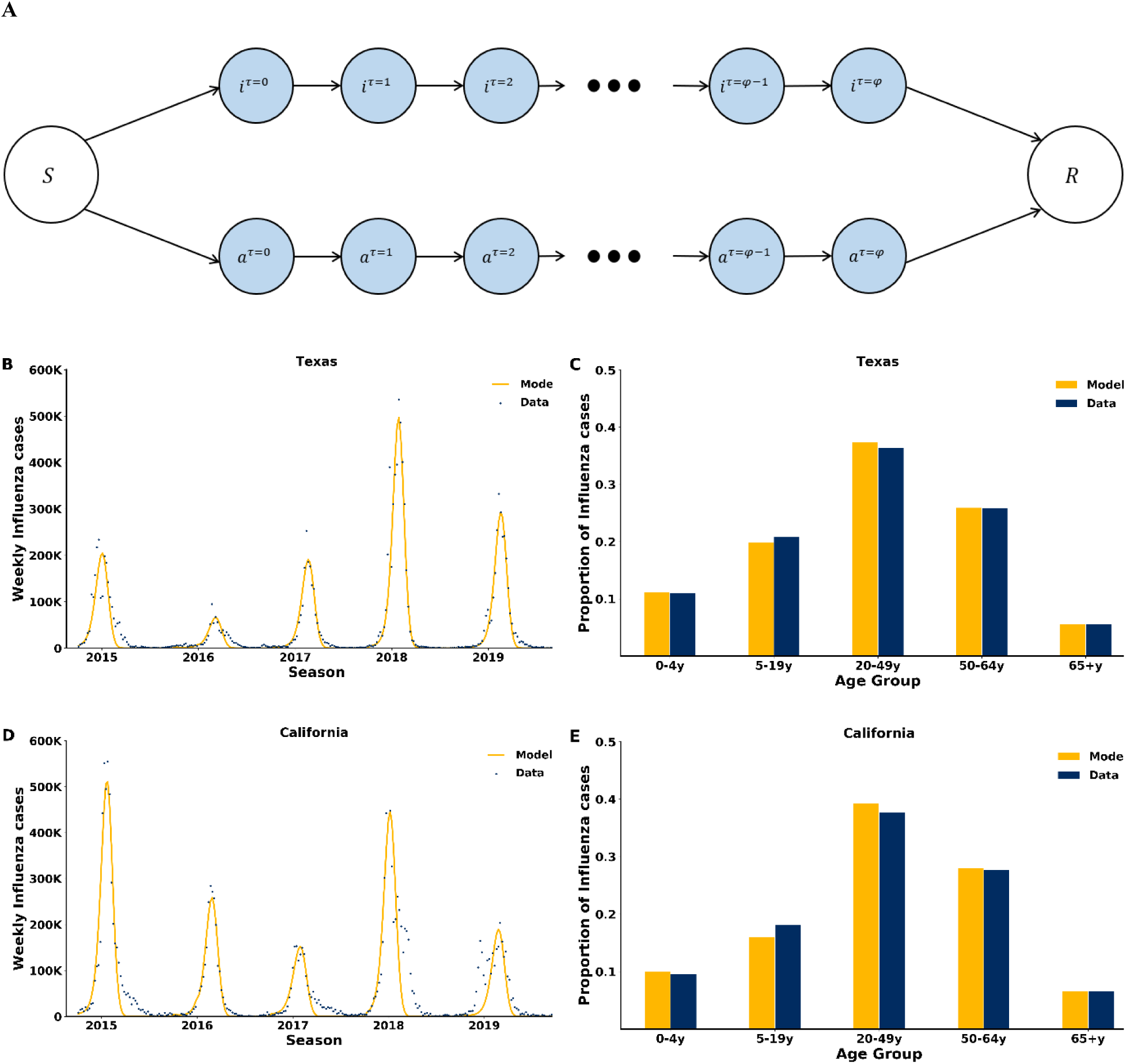
Structure and fit of the model. (A) Compartmental diagram of the transmission model. Following exposure, susceptible individuals S move to the symptomatic or asymptomatic compartment, it=0,at=0, representing the first day of infection. Then, on each following day, they transition to a matching compartment, where they may transmit the disease to others based on 1) their contact mixing patterns, 2) the per-day viral load, 3) the seasonal forcing, and 4) their antiviral treatment regimen. Recovered individuals move to compartment *R*, where they are fully protected for the entire season. For clarity, age and risk stratification are not displayed (*SI Appendix*). (B, D) Time series of recorded weekly influenza cases and model fit to California and Texas (the model fit to Connecticut and Virginia is provided in *SI Appendix*, figure supplementary S2). (C, E) Data and model fit to the age distribution among influenza infections.

Susceptible individuals in the model may interact with infectious individuals and become either asymptomatically or symptomatically infected (Furuya-Kanamori et al., 2016; Leung et al., n.d.), where they can transmit the disease to others until recovery. Consistent with previous models (Medlock and Galvani, 2009; Ndeffo Mbah et al., 2013; Yamin et al., 2014), we assumed that upon recovery, individuals are fully protected for the entire season. This assumption is supported by prospective studies demonstrating that reinfection in the same season is rare (Möst et al., 2019; Möst and Weiss, 2016).

### Force of Infection

The rate at which infectious individuals transmit depends on 1) age-specific contact rates (Table supplementary S1) between an infected individual and his or her contacts, 2) age-specific susceptibility to infection, and 3) infectiousness of the infected individual based on her/his daily viral loads and time in the season (Figure supplementary S1, and SI Appendix for details).

In the US, influenza incidence is seasonal, with a peak typically striking in the winter, yet the driver for this seasonality remains uncertain (Lipsitch and Viboud, 2009). Thus, we included general seasonal variation in the susceptibility rate of the model as 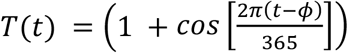, where ϕ is a seasonal offset. This formulation was previously shown to accurately capture the seasonal variation in the incidence of respiratory diseases by US state (Pitzer et al., 2015; Yamin et al., 2016).

### Hospitalizations

Hospitalization was not model explicitly. However, we computer the number of hospitalizations for each age and risk-group by multiplying the number of symptomatic infected individuals by the rate of hospitalization given influenza infection. These age- and risk-specific rates were obtained from epidemiological studies (Pitzer et al., 2015; Yamin et al., 2016).

### Baseline Vaccination and Treatment

For each year, we parameterized vaccination uptake from state-specific influenza vaccine coverage data for different age groups, as observed from 2013 to 2018 (Table supplementary S2) (“2010-11 through 2018-19 Influenza Seasons Vaccination Coverage Trend Report | FluVaxView | Seasonal Influenza (Flu) | CDC,” n.d.). We estimated vaccine efficacy using the CDC estimates for influenza vaccine efficacy between 2013 and 2018 (“CDC Seasonal Flu Vaccine Effectiveness Studies | CDC,” n.d.).

Antiviral treatment is provided to high-risk individuals who seek care in health clinics and hospitals. We parameterized our model using data from recent large-scale studies on the time to seek care and antiviral prescription among laboratory-confirmed high-risk influenza patients in the US (Biggerstaff et al., 2014). These data were used to inform our baseline treated scenario for each state.

High-risk individuals receiving antiviral treatment have a reduced rate of hospitalization compared to nontreated high-risk patients. The age-specific reduction in hospitalization rate among treated high-risk individuals was obtained from previous retrospective and prospective studies (Kaiser et al., 2003; Piedra et al., 2009).

### Model Calibration

We calibrated our model to weekly cases of influenza (confirmed by viral isolation, antigen detection, or PCR) and influenza-like illnesses (ILI) cases reported (“FluView Interactive | CDC,” n.d.) to estimate empirically unknown epidemiological parameters (SI Appendix, Tables supplementary S3 and S4). These data were collected by the CDC’s National Respiratory and Enteric Virus Surveillance System and state health departments from 2014 to 2019. We used data from a recent meta-analysis (Tokars et al., 2018a) of seasonal influenza in the US between 2011 and 2016 to obtain the median annual attack rate per age group.

### Interventions

We evaluated two interventions for increasing the number of high-risk patients seeking care and being treated within the first two days. In the first intervention, we increased the number of individuals who are treated within the first two days of symptoms onset by assuming that a proportion of those who received treatment after the first two days of symptoms onset would receive treatment within the first two days of symptoms onset. In the second intervention, we increased the total number of infected high-risk individuals who received treatment while assuming that they all received treated within the first two days of symptoms onset. We evaluated the population- and individual-level benefits of these interventions in terms of infections and hospitalizations averted in during a single influenza season.

### Sensitivity Analysis

We conducted sensitivity analyses to examine the robustness of our results. In the first analysis, we investigated the impact of effective vaccination coverage on the effectiveness of antiviral treatment. Effective vaccination coverage is defined as the product of vaccine efficacy times vaccine coverage, and represents the level of vaccine induced immunity in the population. In the second analysis, we conducted a two-way sensitivity analysis to investigate the joint impact of changing both the attack rate and the effective vaccination coverage.

## Results

The transmission model was calibrated to age-stratified weekly incidence data for the 2014-2015 to 2018-2019 influenza seasons in four states across the US (Figure 1 and figure supplementary S2). The model explicitly accounted for changes in transmissibility with disease progression and timing of antiviral treatment. The model captures the influenza weekly trends and the age distribution of infected individuals (Figure 1 B-E and SI Appendix). For example, in influenza season 2014-2015, the calibrated model accurately showed that influenza infections peaked in week 14 in Texas and week 17 in California, week 19 in Connecticut and week 20 in Virginia (Figure 1 and figure supplementary S2), and the age distributions of influenza cases were consistent with the empirical data in both states (Figure 1 and figure supplementary S2).

We simulated five influenza seasons and projected the number of cases and influenza-induced hospitalization that would have been averted by early treatment. Specifically, we evaluated the population-level benefit of increasing the proportion of high-risk individuals who initiate treatment within 48 hours of symptoms onset without increasing the baseline number of treated individuals (Figure 2 and figure supplementary S3). We found that keeping the same number of treatments while providing earlier treatment can substantially decrease transmission, thereby reducing both influenza illnesses and hospitalizations. For example, if 60% of high-risk patients who actually received treatment more than 48 hours after symptoms onset were treated earlier, it would avert an additional 58,482 (45,066-74,997) cases and 502 (353-752) hospitalizations annually (Figure 2 A and B) in Texas, 81,472 (66,527-100,836) cases and 743 (579-998) hospitalizations in California (Figure 2 C and D), 6,303 (5,662-9,144) case and 57 (50-90) hospitalizations in Connecticut (Figure supplementary S3 E and F), and 16,380 (13,995-18,386) cases and 140 (117-161) hospitalizations in Virginia (Figure supplementary S3 G and H). If all patients were treated within 48 hours of symptoms onset, it would avert an additional 65,210 (50,206-83,713) cases and 543 (382-811) hospitalizations annually in Texas, 90,842 (74,126-112,531) cases and 805 (627-1,080) hospitalizations in California, 7,012 (6,297-10,187) cases and 62 (54-98) hospitalizations in Connecticut, and 18,230 (15,567-20,471) cases and 152 (127 - 175) hospitalizations in Virginia (Figure 2 and figure supplementary S3).

**Figure 2.**
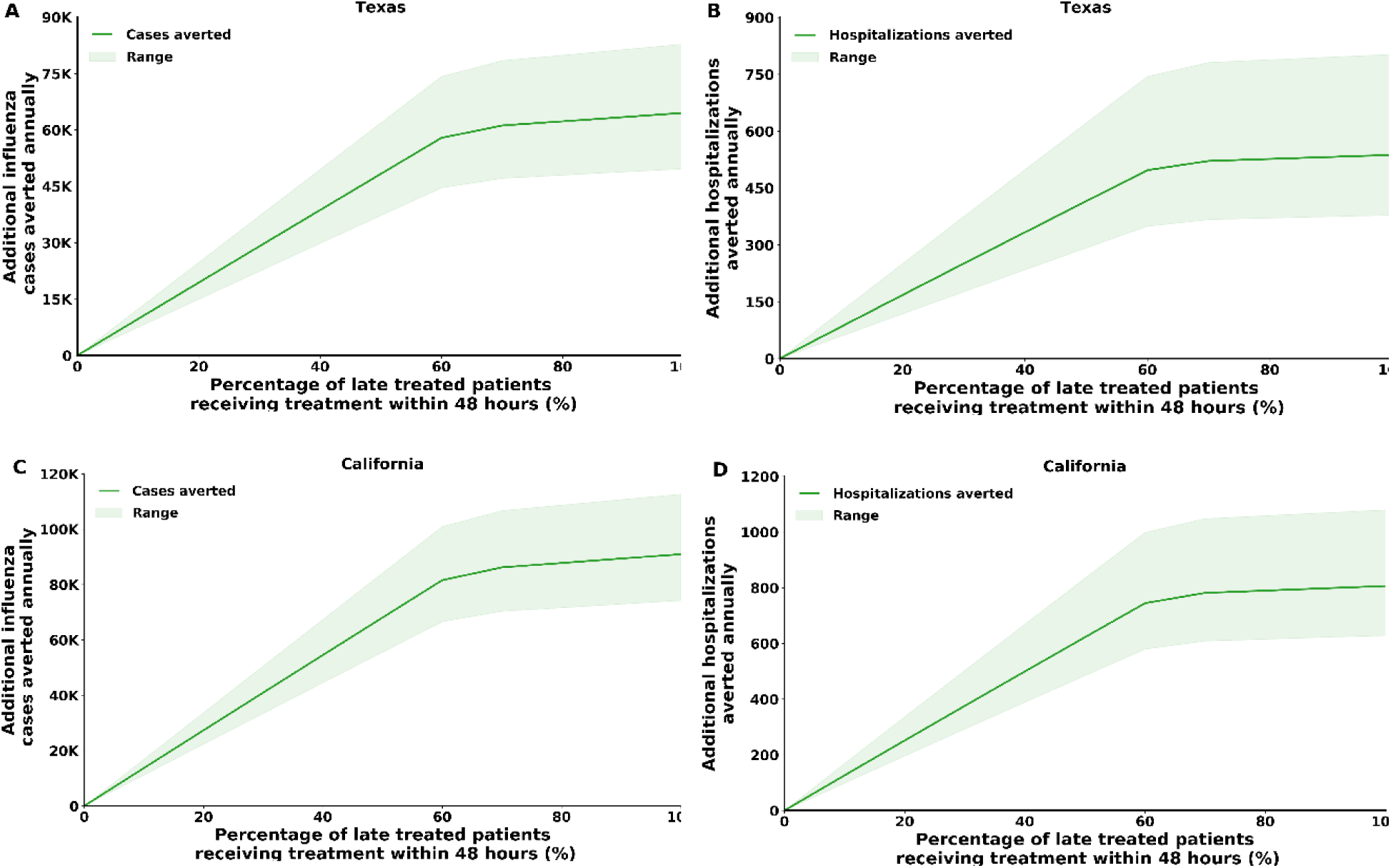
Model projection of additional influenza cases and hospitalizations averted annually in California and Texas by increasing the number of high-risk patients who initiate treatment within 48 hours of symptoms onset. Here, a proportion of high-risk patients who initiated antiviral treatment more than 48 hours after symptoms onset was assumed to receive treatment earlier. (A, C) Total number of infection cases averted. (B, D) Total number of hospitalizations averted.

We also evaluated the benefit of increasing antiviral treatment coverage among high-risk individuals. Increasing early treatment coverage among individuals at high risk has a substantial benefit in terms of averting both cases and hospitalizations (Figure 3 and figure supplementary S4). For example, if 20% of high-risk individuals infected with influenza were treated within 48 hours of symptoms onset, it would avert, on average, 115,641 influenza cases annually in Texas, 160,645 cases in California, 12,571 cases in Connecticut, and 32,494 cases in Virginia. In addition to cases averted, this increase in treatment coverage would avert, on average, 1,068 hospitalizations annually in Texas, 1,587 hospitalizations in California, 121 hospitalizations in Connecticut, and 295 hospitalizations in Virginia.

**Figure 3.**
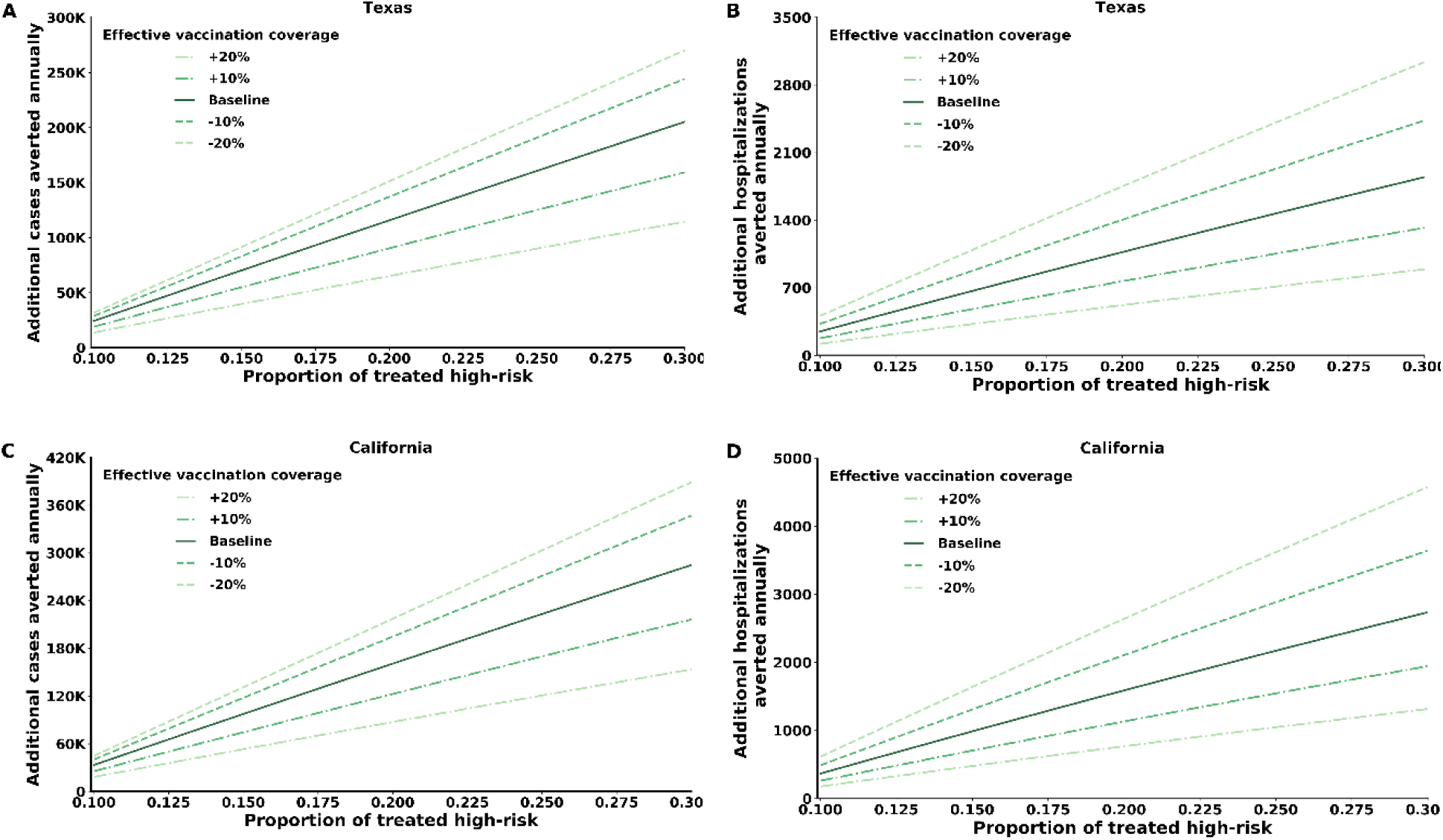
Model projections of influenza cases and hospitalizations averted in California and Texas by increasing the portion of treated high-risk individuals who seek care and receive treatment within 48 hours of symptoms onset. For the sensitivity analysis, we increased and decreased the vaccination coverage by 10 and 20 percent. (A, C) Total number of cases averted. (B, D) Total number of hospitalizations averted.

Given that the benefit of treatment depends on the underlying vaccination coverage within the population, we also examined how variation in vaccination coverage affects the benefit of treating high-risk individuals. We found that increasing effective vaccination coverage would decrease both influenza burden and the benefit of treatment (Figure 3). Nevertheless, for a 20% increase in influenza effective vaccination coverage among all age groups, our results suggest that the benefit of treatment remains substantial (Figure 3). For example, treating 20% of infected high-risk individuals within 48 hours of symptoms onset would avert 65,018 cases and 519 hospitalizations in Texas (Figure 3 A and C). In California, it would avert 87,243 cases and 766 hospitalizations (Figure 3 B and D). In Connecticut, it would avert 4,621 cases and 40 hospitalizations, and in Virginia it would avert 12,347 cases and 100 hospitalizations (Figure supplementary S4).

To estimate the benefits of a policy that targets specific age groups for early treatment, we evaluated the effectiveness of age-targeted treatment strategies for averting influenza cases and influenza-induced hospitalizations (Figure 4 and figure supplementary S5). We found that treatment of the elderly (>65 years old) has the highest impact on reducing hospitalizations (Figure 4 B and D). This result was driven mainly by the fact that this age group has the highest risk for influenza complications, which leads to a higher rate of hospitalization. The highest impact on reducing transmission was achieved by targeting high-risk individuals aged 5-19 years old. For example, in Texas, early treatment of the 5-19 years old age group will avert 2.31 cases per treatment, and early treatment of the >65 years old age group will avert 0.04 hospitalizations per treatment.

**Figure 4.**
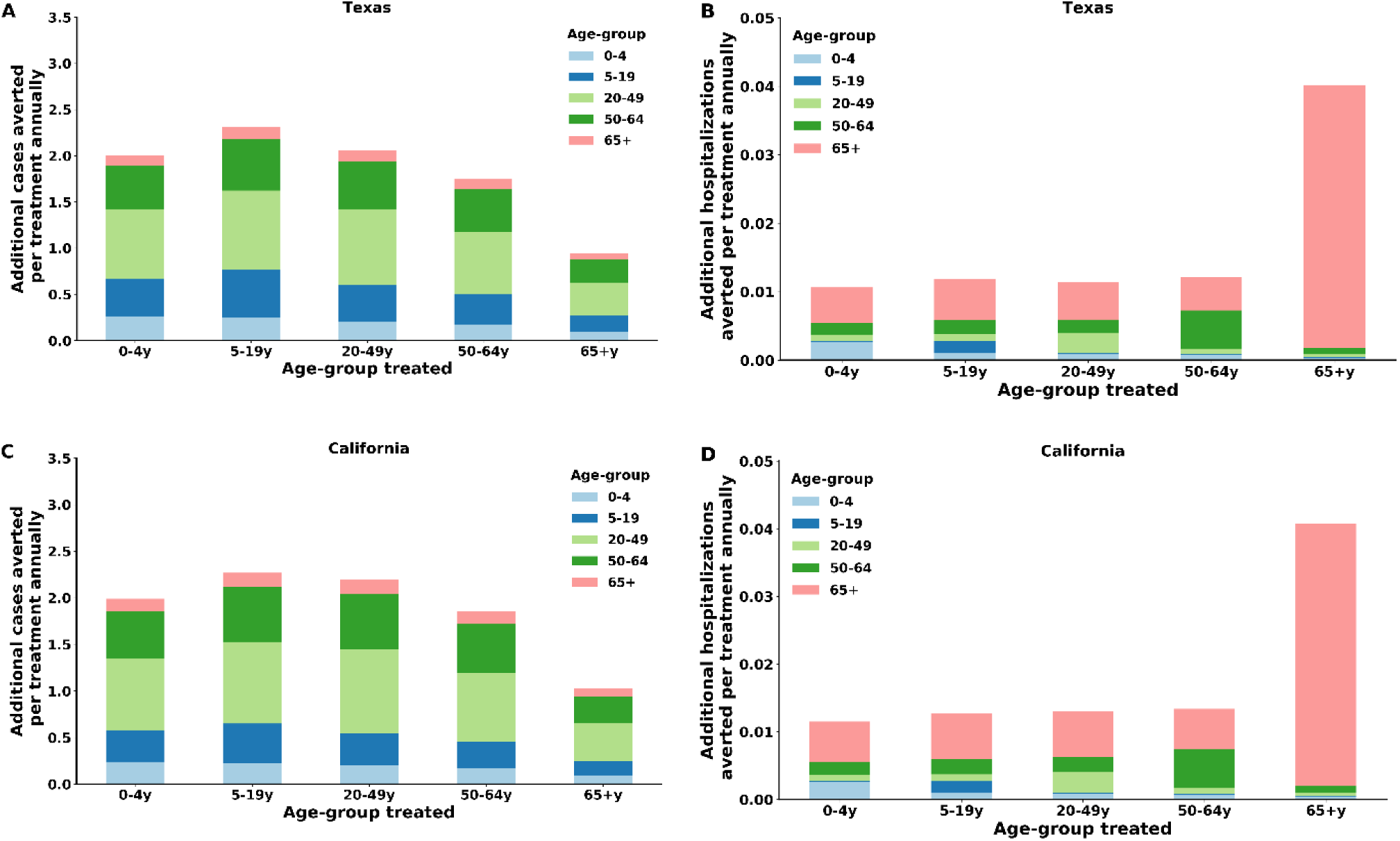
Model projections of influenza cases and hospitalizations averted per treatment by treating each age group within 48 hours of symptoms onset in California and Texas. (A, C) Number of cases averted per treatment for each age group stratified by age group. (B, D) Number of hospitalizations averted per treatment for each age group stratified by age group.

The yearly attack rate of influenza varies considerably between seasons. Thus, we explored the benefit of treating high-risk patients under different attack rates and effective vaccination coverage. We found that the benefit conferred by treatment decreases with increasing attack rate and increases with increasing effective vaccination coverage (Figure 5 and figure supplementary S6). With a medium attack rate of 12.03% and 11.86% effective vaccination coverage, which is 60% from the yearly mean effective vaccination coverage, 0.69-0.78 cases are averted per treatment in California. With an attack rate of 5.70% and the same effective vaccination coverage, 0.96-1.11 cases are averted per treatment. For the same transmission settings in Texas, for a yearly attack rate of 15% and 12.11% effective vaccination coverage, 0.49-0.55 cases are averted per treatment. With an attack rate of 2.81% and the same effective vaccination coverage, 1.65-1.81 cases are averted per treatment. In Texas, the benefit conferred by treatment was found to decrease when the effective vaccination coverage exceeds 22.50% (Figure 5C). In all settings considered, the marginal benefit of treatment per dose was found to decrease with increasing treatment coverage and vaccination coverage (Figure 5). This saturation in the benefit of treatment is driven by the increased herd immunity resulting in a decrease in the indirect benefit of treatment.

**Figure 5.**
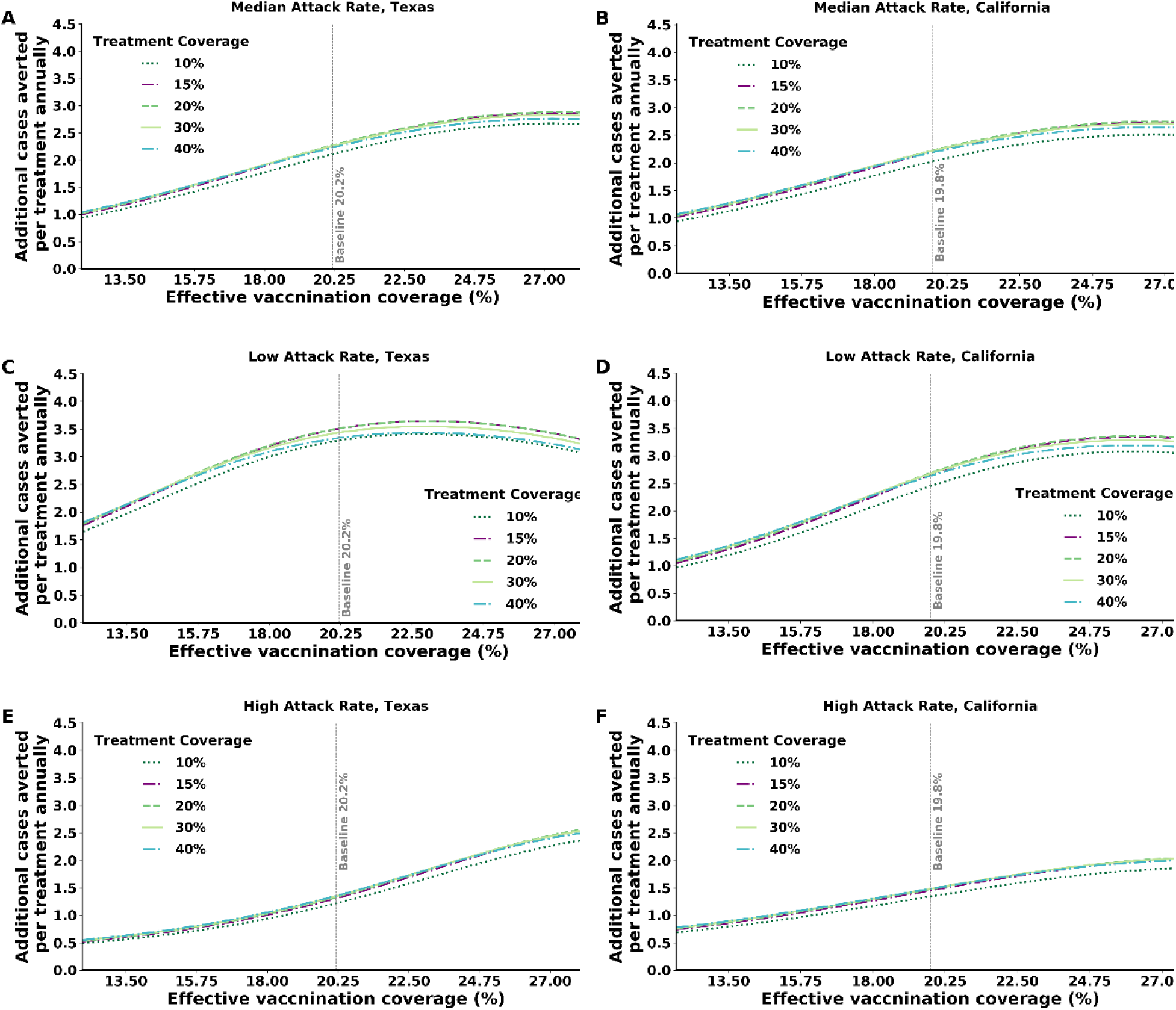
The mutual effect of attack rate and vaccination coverage in California and Texas on the number of cases averted per treatment for each treatment coverage among high-risk individuals infected with influenza. Infected high-risk individuals seek care and receive treatment within 48 hours of symptoms onset. (A, B) Median attack rate settings. (C, D) Low attack rate settings. (E, F) High attack rate settings.

## Discussion

Our key finding shows that increasing the timeliness of treatment of high-risk patients, even without increasing the current treatment coverage, is highly effective in reducing morbidity and mortality associated with influenza at the population level. The reason behind this finding is that the viral load of influenza is the highest during the first three days from symptoms onset. Earlier treatment reduces the viral loads and thus has a pivotal nonlinear effect of reducing transmission.

Interventions that could improve the timeliness of high-risk patients seeking care and their access to timely antiviral prescriptions and potentially reduce influenza-associated morbidity and mortality are urgently needed. These interventions could include education of high-risk patients, education of physicians about the benefits of early antiviral treatment among high-risk patients, and innovative tools to enhance early detection of influenza infection and treatment. These tools include providing phone consultations or remote electronic visits (virtual visits).

Vaccination remains the main tool for controlling seasonal influenza. However, vaccine efficacy varies widely between influenza seasons, and vaccination coverage remains suboptimal (“CDC Seasonal Flu Vaccine Effectiveness Studies | CDC,” n.d.; Doyle et al., 2019). Our study shows that the benefit of treating early and increasing the treatment coverage is substantial, regardless of vaccine efficacy and coverage. Counterintuitively, we found that the higher the effective vaccine coverage is, the higher the marginal outcome of treatment. This phenomenon is driven by the fact that high effective vaccination coverage results in low disease transmission, which in turn increases the indirect benefit of treatment. This finding emphasizes the importance of antiviral treatment as a complementary effort to vaccination.

Despite the effectiveness of antivirals in reducing influenza-related morbidity and mortality, the emergence of drug resistance poses a critical limitation on their application. Therefore, parsimonious use of antivirals is needed to mitigate the emergence of influenza antiviral-resistant strains. Studies have suggested that to reduce the risk of antiviral overuse while maximizing their use to mitigate the burden of influenza, low-risk patients should be tested before treatment with antivirals and high-risk patients with clinically diagnosed influenza infection should receive prompt treatment pending results of a laboratory-confirmed test (Sintchenko et al., 2002). Our study shows that increasing the timeliness of treatment without increasing the number of treated individuals would substantially increase the population-level benefit of antiviral treatment. For example, if the proportion of high-risk patients who receive treatment within the first 48 hours of symptoms onset were to increase from its baseline value of 8.1% to 14.85% (the total number of high-risk patients who receive treatment: both within and after 48 hours), it could avert an additional 65,210 (50,206-83,713) cases annually in Texas, 90,842 (74,126-112,531) cases in California, 7,012 (6,297-10,187) cases in Connecticut, and 18,230 (15,567-20,471) cases in Virginia.

The ongoing coronavirus (COVID-19) pandemic has already put unprecedented strain on the health system of many countries. As the disease continues to unfold across the world, its impact on national health systems is yet to be fully understood. In the US, the possibility of COVID-19 transmission during the next influenza season is raising a substantial concern about the health system being overwhelmed by visits from both COVID-19 and influenza-related complications among high-risk patients. The specter of this challenging scenario emphasizes the importance of the results of this study and the urgent need for increase influenza vaccine uptake and timeliness of antiviral treatment among high-risk patients in the US

Our study includes several limitations that should be addressed by future studies. Although several studies have attempted to estimate the annual attack rate of influenza in the US (Jayasundara et al., 2014; Molinari et al., 2007; Tokars et al., 2018b), the state-level rates remain unknown. Therefore, we used the nationwide attack rate to normalize the state-specific influenza cases. Moreover, we used nationwide data to estimate the treatment coverage and timeliness for each state, as state-specific data are not available. Under these assumptions, our results were qualitatively similar across all states, with quantitative differences being driven by state-specific information on population size and demography, vaccination coverage, and influenza seasonality.

In conclusion, increasing the timeliness and coverage of antiviral treatment among high-risk individuals has the potential to substantially reduce the burden of seasonal influenza in the US. Timely treatment not only reduces the risk of influenza-induced hospitalization for the treated individual but may also reduce disease transmission. Public health decision makers should invest continuous efforts to follow the CDC guidelines by treating influenza patients at high risk as soon as possible.

## Data Availability

Medical data are publicly available.

## Acknowledgement

None

## Supplementary material for: Optimizing antiviral treatment for seasonal influenza in the United States

## 1. Model

### 1.1. The model

We developed a dynamic model for age-stratified Influenza infection progression and transmission in four states in the United States. Our model is a modified Susceptible-Exposed-Infected-Recovered (SEIR) compartmental framework (Vynnycky and White, 2010), whereby the population is stratified into health-related compartments, and transitions between the compartments occurs over time (Main text, Figure 1). To model age-dependent transmission, we stratified the population into n = 5 age groups: 0–4 years, 5-19 years, 20-49 years, 50-64 years, and ≥ 65 years. Consistent with immunological observations (“Antibodies Cross-Reactive to Influenza A (H3N2) Variant Virus and Impact of 2010–11 Seasonal Influenza Vaccine on Cross-Reactive Antibodies — United States,” n.d.; Branch et al., 2012; Hancock et al., 2009; Mandelboim et al., 2014; Ranjeva et al., 2019; Sharabi et al., 2016), we assumed age-dependent susceptible reduction due to preexisting serum influenza neutralizing antibodies from previous exposure which reduce susceptibility to infection. Consistent with previous models (Medlock and Galvani, 2009; Ndeffo Mbah et al., 2013; Yamin et al., 2014), we assumed that upon recovery individuals are fully protected for the entire season. This assumption is also supported by prospective studies demonstrating that reinfection in the same season is rare, yet possible (Möst et al., 2019; Möst and Weiss, 2016).

Accordingly, we stratified the population into four health-related compartments: susceptible *S*_*j,k*_(*t*), symptomatic infectious *I*_*j,k*_(*t*), asymptomatic infectious *A*_*j,k*_(*t*), and recovered *R*_*j,k*_(*t*), such that at any given time t (in days):

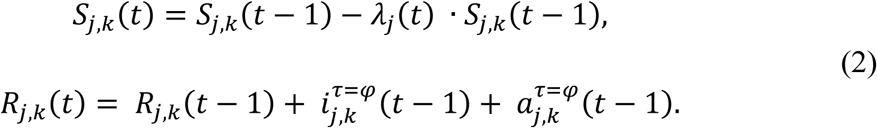

where the index *j* ∈ {1,2, …, *n*} represents the age-group of each individual, and the index *k* ∈ {*L, H*} specifies the risk-group of each individual (i.e. High-risk, or low-risk).

### 1.2. Model transitioning

Before the beginning of each influenza season, individuals start from susceptible compartment *S*_*j,k*_(0). Individuals who are immune, due to preexisting serum influenza neutralizing antibodies from previous exposure (proportion of *ξ*_*j*_ of each age-group), are not included in the susceptible compartment. Thus, they are transitioned to the recovered compartment *R*_*j,k*_(0) according to their age-group and risk-group. Susceptible individuals can get vaccinated with vaccination coverage *v*_*j,k*_ with the efficacy of the vaccine η. Individuals who are effectively vaccinated transitioned to the *R*_*j,k*_(0) compartment, where they are fully protected for the entire season. Susceptible individuals can become infected with a force of infection *λ*_*j*_(*t*), depending on their age group j. Infectious individuals can be asymptomatic with the probability of *f* and symptomatic with probability of (1 − *f*). Infected individuals remain in the infectious compartments *I*_*j,k*_(*t*), *A*_*j,k*_(*t*) for φ days and can infect their contacts based on their daily viral load and daily contact behavior (SI Appendix 2.1 Data set and parameters). After infectious period, individuals’ transition into the recovered compartment *R*_*j,k*_(*t*), and remain recovered until the end of the season. In the case of asymptomatic infection or treated high-risk, contact mixing is unaffected, but transmission is lower than during symptomatic infection due to a lower viral load (SI Appendix, 2.1 Data set and parameters).

To incorporate the evolution of infectiousness during infection, we explicitly track the number of symptomatic and asymptomatic infected individuals 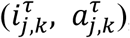, respectively, with regard to their day of infection, τ. Hence, 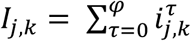 and 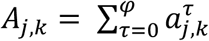. Thus, the transmission model is composed of the following system of difference equations:

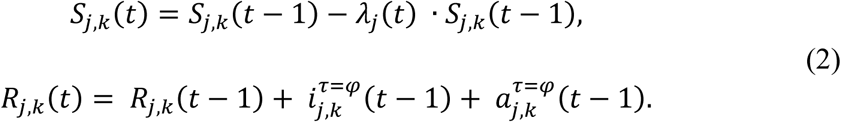

with daily numbers of infected individuals:

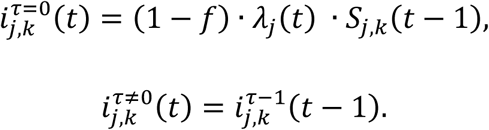

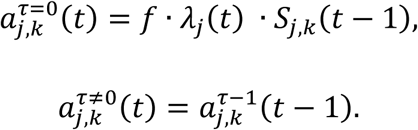

With initial conditions:

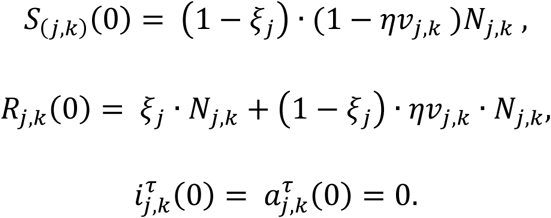

### 1.3. Force of infection

The rate at which individuals transmit Influenza at time t is *λ*_*j*_(*t*). This rate depends on the combination of 1) age-specific contact rates between an infected individual and his or her contacts, 2) infectiousness of the infected individual based on his or her daily viral loads, and 3) age-specific susceptibility to infection. In the US, influenza incidence is seasonal, with a peak typically striking in the winter, yet the driver for this seasonality remains uncertain(Lipsitch and Viboud, 2009). Thus, we included general seasonal variation in the susceptibility rate of the model as

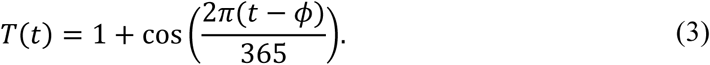

The seasonal offset *ϕ* will be calibrated, for each season separately, in order to fit the influenza case data. We set the boundaries of the search between the fourth week and the twenty-seventh week of the season (according to the data). This formulation was previously shown to accurately capture the seasonal variation in the incidence of respiratory diseases by US state (Pitzer et al., 2015; Yamin et al., 2016).

We incorporate age-specific contact patterns between individuals, represented by contact rate between an infected individual in age-group e and each of their contacts with susceptible in age-group j, denote by *C*_*e,j*_. i.e. the contact matrix C will be detailed in the next section.

For high-risk individuals from age-group *j*, we parameterize the timing of the antiviral treatment uptake (see SI Appendix 2.1 Data set and parameters). For each day during the exposed and infection periods (τ), we incorporate the proportion of untreated high-risk individuals 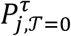, the proportion of high-risk individuals who got treated on the second day since symptoms onset 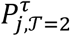, and the proportion of high-risk individuals who got treated on the third day since symptoms onset 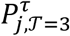. high-risk individuals who got treated after the third day since symptoms onset are considered as treated ineffectively, with treatment having no impact on disease progression and severity (Aoki et al., 2003; Heinonen et al., 2010; “Use of Antivirals | CDC,” n.d.). Therefore, we include them in 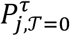.

Given a contact with an infected host, the logarithm of the infectious viral load has been shown to be correlated with the transmissibility of several respiratory viruses(Couch et al., 1969; Tellier, 2009). The logarithm of the viral load depends on the risk-group of the infected individual *k* ∈ {*H, L*}, the timing of the antiviral treatment for the high-risk individuals T, the day of infection *τ* which includes the exposed and infection periods, and on the type of infection. (see SI Appendix 2.1 Data set and parameters). In addition, we consider the age-specific susceptibility rate of and individual *β*_*j*_, was parametrized by calibrating our model with weekly influenza records (See Section 2.2. calibrated parameters). Taken together, the force of infection *λ*_*j*_(*t*) is given by:

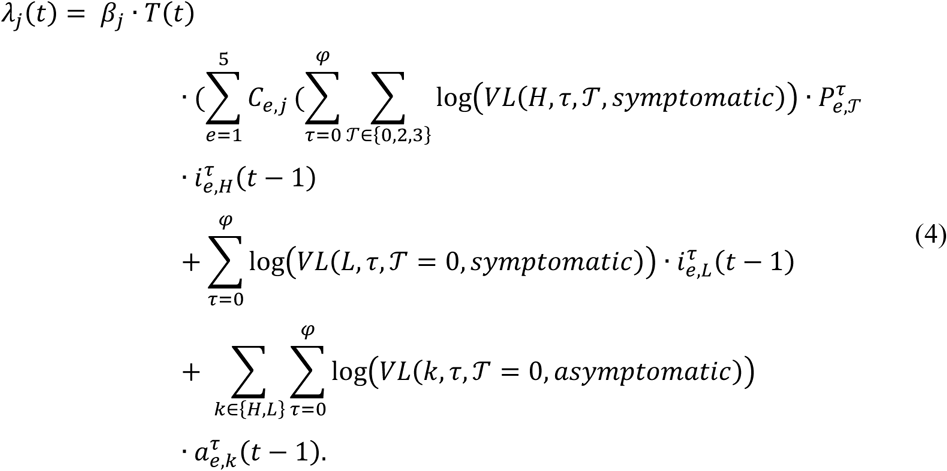

## 2. Data set and parameters

### 2.1. Fixed parameters

#### Contact mixing

We parameterized the age-specific contact rates between an infected individual e and their contact *j, C*_*e,j*_ based on the contact matrix parameterized by a previous study(Yamin et al., 2016). We adjusted the contact mixing matrix to our model age-groups. For age-groups, 5 − 19y, 20 − 49y we analyzed the data used to build the contact matrix using the same methods described in that study. This contact data exhibits frequent mixing between similar age-groups, moderate mixing between children and adults in their thirties (likely their parents), and infrequent mixing between other groups.

**Table S1.** Age-specific rates *C*_*e,j*_ between an infected individual e and their contact *j*.

|  |  |  |  |  |  |
| --- | --- | --- | --- | --- | --- |
| 0-4 years | 43 | 18 | 11 | 6 | 3.8 |
| 5-19 years | 18 | 37 | 10 | 8.9 | 3.9 |
| 20-49 years | 11 | 10 | 24 | 14 | 6 |
| 50-64 years | 6 | 8.9 | 14 | 18 | 8.1 |
| ≥ 65 years | 3.8 | 3.9 | 6 | 8.1 | 12 |
|  | 0-4 years | 5-19 years | 20-49 years | 50-64 years | ≥ 65 years |

#### Viral load

Using recent prospective studies of the course of Influenza infections in young children and adults, we estimated the viral load for asymptomatic, symptomatic high- and low-risk and treated high-risk who got treated on the first three days since symptoms onset (Ip et al., 2017; Lee et al., 2013).

Viral load for asymptomatic estimated using the results of a prospective study that tracked after households in Hong Kong with an influenza confirmed patient (Ip et al., 2017). To estimate the viral load for the asymptomatic infected individuals, we smoothed the data of paucisymptomatic.

For symptomatic high- and low-risk and treated high-risk who got treated on the first three days from symptoms onset, we estimated the viral load using the data of a prospective study monitored patients with laboratory-confirmed influenza who admitted to the medical department of the Prince of Whales Hospital(Lee et al., 2013). Day zero and days eight to fourteen since symptoms was estimated by fitting an exponential curve to the data. To estimate the viral load during the exposure period we scaled the data using the data of asymptomatic. For treated high-risk, the viral load prior to initiating treatment was set to be the same as for not treated high-risk individuals (Figure S1).

**Figure S1.**
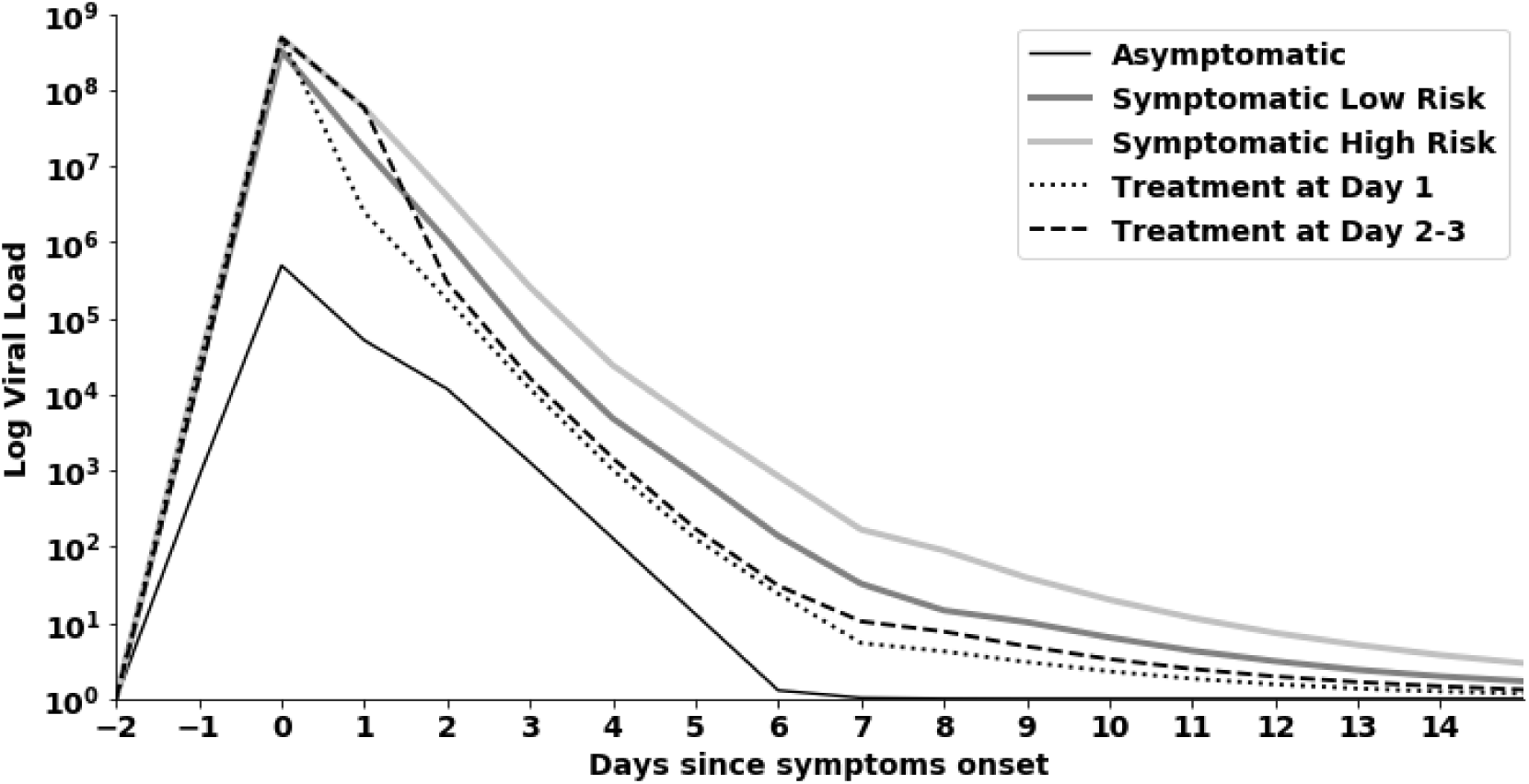
Daily log viral load following influenza infection for asymptomatic, symptomatic high and low-risk and treated high risk who got treated on the first three days since symptoms onset.

#### Hospitalizations

The number of hospitalizations for each age- and risk-group was calculated by multiplying the number of symptomatic infected individuals by the probability of hospitalization given influenza infection (See Table S3). Using data from epidemiological study, we calculated the fraction of symptomatically infected individuals that will be hospitalized based on the ratio between hospitalizations and infection cases stratified by age (Rolfes et al., 2018). For age-groups 5 − 19y, 20 − 49y, 50 − 64y, ≥ 65y, the proportion of hospitalizations related to each risk-group estimated based on the ratio between hospitalizations of high-risk and low-risk as suggested by previous epidemiological study(Mullooly et al., 2007). For age-group 0 − 4y a previous retrospective study suggests that 37% of hospitalizations related to high-risk children (Ampofo et al., 2006). These data are consistent with the US Influenza hospitalization surveillance network data (Chaves et al., 2015).

Infected high-risk individuals who treated within three days from symptoms onset have a lower probability to be hospitalized (Kaiser et al., 2003; Piedra et al., 2009). For individuals who younger than 19 years old, we reduced the probability of hospitalization given an infection by 75%(Piedra et al., 2009). For adults (older than 19 years old) we reduced the probability by 59%(Kaiser et al., 2003).

#### Vaccinations coverage

We estimated the vaccination coverage based on data obtained from Center for Disease Control and Prevention (CDC) (“2010-11 through 2018-19 Influenza Seasons Vaccination Coverage Trend Report | FluVaxView | Seasonal Influenza (Flu) | CDC,” n.d.). The data is stratified by state, age, season, and risk-group. To adjust the age stratification in the data to the age-groups used in our model, we assumed uniform distribution for each age-group in the data. For age-groups without risk-group stratification we assumed equal vaccination coverage for both low- and high-risk individuals.

We parametrize the vaccination coverage for each year at both national and state-level as observed from 2013-2018 (See Table S2). For the national coverage we used the median vaccination coverage for each year. As some of the states do not have data for season 2013-14, we used the average coverage of seasons 2012-13,2014-15 as the coverage for season 2013-14. In both levels, we used the mean coverage over the five seasons as our baseline.

As mentioned in the equation (2), the vaccination coverage *v*_*j,k*_ is multiplied by the vaccination efficacy *η* to calculate the effective vaccination coverage.

**Table S2.** Average Vaccination coverage (“2010-11 through 2018-19 Influenza Seasons Vaccination Coverage Trend Report | FluVaxView | Seasonal Influenza (Flu) | CDC,” n.d.).

| Age-group | 0-4 years |  | 5-19 years |  | 20-49 years |  | 50-64 years |  | ≥ 65 years |  |
| --- | --- | --- | --- | --- | --- | --- | --- | --- | --- | --- |
| Risk-group | high | low | high | low | high | low | high | low | high | low |
| **US** | 70.08% | 70.08% | 55.92% | 55.92% | 45.90% | 33.99% | 45.90% | 33.99% | 64.56% | 64.56% |
| **California** | 71.26% | 71.26% | 57.44% | 57.44% | 44.45% | 32.20% | 44.45% | 32.20% | 63.14% | 63.14% |
| **Connecticut** | 85.60% | 85.60% | 64.97% | 64.97% | 52.02% | 37.62% | 52.02% | 37.62% | 66.80% | 66.80% |
| **Virginia** | 75.58% | 75.58% | 59.10% | 59.10% | 48.54% | 39.92% | 48.54% | 39.92% | 67.72% | 67.72% |
| **Texas** | 71.60% | 71.60% | 57.87% | 57.87% | 45.38% | 31.60% | 45.38% | 31.60% | 63.10% | 63.10 |

#### Treatment coverage

Antiviral treatment is only provided to high-risk individuals that seek care in health clinics and hospitals. We assumed that following clinic or hospital visit and treatment prescription, it takes at least one day to a patient to initiate course of treatment. Therefore, the earliest treatment initiation time is two days from symptoms onset. Moreover, people who sought care on the third and fourth day since symptoms onset are considered as treated ineffectively, with treatment having no impact on disease progression and severity (Aoki et al., 2003; Heinonen et al., 2010; “Use of Antivirals | CDC,” n.d.). Thus, we included them with the untreated individuals.

Hence, the probability of infected high-risk to seek care and get treated effectively (getting treated within 3 days from symptoms onset) is given by equation (5):

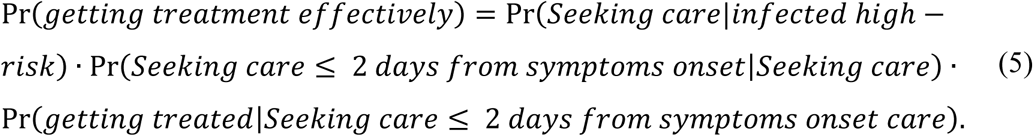

The probability of seeking care given a high-risk infection stratified by age was calculated using data from an epidemiological study (Molinari et al., 2007) (See Table S3).

Using data from recent large scale studies on the time to seek for care and antiviral prescription among laboratory-confirmed high-risk influenza patients in the US (Biggerstaff et al., 2014; Stewart et al., 2018), we estimated the probability of seeking care withing two days from symptoms onset given an infected high-risk who sought for care and the probability of getting treated given an infected high-risk who sought for care withing two days since symptoms onset. Those studies provided age-stratified data on the proportion of treated individuals who sought care during the first two days, 3 to 7 days, and 7 days since symptoms onset.

Further to our assumptions, we assumed high-risk individuals who sought care during the first two days since symptoms onset, have equal probability of treatment initiation time on the second and third day since symptoms onset. High-risk patients treated effectively are accounted for 54.5% of the treated high-risk patients (Biggerstaff et al., 2014).

**Table S3:** Fixed parameters used in the transmission model.

| Parameter | Description | Value | Justification |
| --- | --- | --- | --- |
| $\xi_j$ | Proportion of individuals in age-group $j$ who are immune due to preexisting serum influenza neutralizing antibodies from previous exposure. | 0-4: 0<br>5-19: 0.2<br>20-49: 0.3<br>50-64: 0.25<br>65+: 0.25 | (Mandelboim et al., 2014)<br>(Branch et al., 2012)<br>("Antibodies Cross-Reactive to Influenza A (H3N2) Variant Virus and Impact of 2010–11 Seasonal Influenza Vaccine on Cross-Reactive Antibodies — United States," n.d.)<br>(Hancock et al., 2009)<br>(Sharabi et al., 2016) |
| $N_{j,k}$ | Population size of risk-group $k$ age-group $j$. | Varies between states | (Molinari et al., 2007)<br>("Population Distribution by Age \| The Henry J. |
|  |  |  | Kaiser Family Foundation,” n.d.) |
| $\eta$ | Influenza vaccination efficacy. | 0.45 | (“CDC Seasonal Flu Vaccine Effectiveness Studies \| CDC,” n.d.)<br>(Doyle et al., 2019) |
| $h_{j,k}$ | Probability of hospitalization given an infected individual in age-group $j$ and risk-group $k$. | k=L<br>0-4: 0.0043<br>5-19:<br>0.0003<br>20-49:<br>0.00062<br>50-64:<br>0.0014<br>65+:<br>0.0235<br>k=H<br>0-4: 0.0025<br>5-19:<br>0.0024<br>20-49:<br>0.005<br>50-64:<br>0.0095<br>65+: 0.07 | (Rolfes et al., 2018)<br>(Mullooly et al., 2007)<br>(Ampofo et al., 2006)<br>(Chaves et al., 2015) |
| $f$ | Probability of becoming asymptomatic given an infection. | 0.191 | (Leung et al., n.d.) |
|  |  |  | (Furuya-Kanamori et al., 2016) |
| $\phi$ | Length of Infection since exposure. | 14 days | (Bell et al., 2006) |
| $P_{j,\mathcal{T}}^{\tau}$ | Proportion of treated high-risk in age-group $j$ on day $\tau$, initiate treatment on day $\mathcal{T} \in \{0,2,3\}$ from symptoms onset. $\mathcal{T} = 0$ means either no treatment provided or treatment ineffectively. | $\tau \geq 2$<br>$\mathcal{T} = 2 \text{ or } 3$<br>0-4: 0.086<br>5-19: 0.054<br>20-49: 0.095<br>50-64: 0.074<br>65+: 0.063 | (Molinari et al., 2007)<br>(Biggerstaff et al., 2014)<br>(Stewart et al., 2018) |
| $C_{e,j}$ | contact rate between an infected individual in age-group $e$ and each of their contacts with susceptible in age-group $j$. | | (Yamin et al., 2016) |
| VL (k, $\tau$, $\mathcal{T}$, asymptomatic /symptomatic) | Viral load of infected individuals in risk-group $k$, on day of infection $\tau$ given treatment initiation day and symptomatic or asymptomatic illness. | | (Ip et al., 2017)<br>(Lee et al., 2013) |

### 2.2. Calibrated parameters

#### Influenza cases data and case definition

To estimate empirically unknown epidemiological parameters, we calibrated our model to the weekly number of influenza incidence according to the CDC data. We obtained the weekly ILI cases for each state and season. We also obtained the weekly proportion of positive specimen to influenza (confirmed by viral isolation, antigen detection, or PCR). These data were collected by the CDC’s National Respiratory and Enteric Virus Surveillance System and state health departments during 2014 to 2019 (“FluView Interactive | CDC,” n.d.). To account for the influenza cases, we multiply the weekly ILI cases by the proportion of positive specimens to influenza. Moreover, to derive the weekly cases for each age-group, we assumed that the proportion of weekly cases for each age-group is the same as the state’s population age structure. The age-group distribution for each state was obtained from the Henry J. Kaiser Foundation’s database (“Population Distribution by Age | The Henry J. Kaiser Family Foundation,” n.d.).

To account for unreported cases, the influenza weekly cases data described above was adjusted to fit the median attack rate of the age-group according to Yamin et. al (Yamin et al., 2016). We used data from a recent meta-analysis study, of seasonal influenza in the US between 2011-2016, to obtain the median annual attack rate per age-group (Tokars et al., 2018). To derive the influenza weekly incidence, we divided the weekly influenza cases by the mean of the seasonal cases such that the yearly average attack rate is equal to the median attack rate.

#### Calibration

To calibrate the model to the incidence data we minimized the squared error between model predictions and incidence data. This is equivalent to maximum likelihood estimation assuming a normal distribution of the error. We conducted this calibration for each of the five seasons (2014-2019) separately. For the calibration, we assumed the median US vaccination coverage by age to account for the variation in attack rates due to vaccination uptake. Also, we assumed the same susceptibility rate for both low- and high-risk in each age-group.

The final transmission model (Main text Figure 1A) included five parameters to be estimated through model calibration: seasonal offset *ϕ*; seasonal susceptibility rate *β*_*j*_ for age-group *j*: 0-4y, 5-49y, 50-64y, 65y.

**Table S4.** Calibrated parameters.

| State | Season | Seasonal<br>offset<br>$\phi$ | Susceptibility<br>among age-group 0-4y<br>$\beta_{0-4}$ | Susceptibility<br>among age-group 5-49y<br>$\beta_{5-49}$ | Susceptibility<br>among age-group 50-64y<br>$\beta_{50-64}$ | Susceptibility<br>among age-group 65+y<br>$\beta_{65+}$ |
| --- | --- | --- | --- | --- | --- | --- |
| California | 2014-15 | 68.724 | 0.0032 | 0.0015 | 0.0031 | 0.0021 |
|  | 2015-16 | 90.950 | 0.0031 | 0.0015 | 0.0030 | 0.0021 |
|  | 2016-17 | 67.791 | 0.0028 | 0.0014 | 0.0028 | 0.0020 |
|  | 2017-18 | 47.705 | 0.0031 | 0.0015 | 0.0030 | 0.0021 |
|  | 2018-19 | 92.125 | 0.0028 | 0.0015 | 0.0029 | 0.0021 |
| Texas | 2014-15 | 44.683 | 0.0026 | 0.0015 | 0.0029 | 0.0021 |
|  | 2015-16 | 97.567 | 0.0026 | 0.0015 | 0.0028 | 0.0022 |
|  | 2016-17 | 87.091 | 0.0028 | 0.0015 | 0.0029 | 0.0021 |
|  | 2017-18 | 73.079 | 0.0029 | 0.0016 | 0.0032 | 0.0022 |
|  | 2018-19 | 88.573 | 0.0028 | 0.0015 | 0.0031 | 0.0022 |
| Connecticut | 2014-15 | 82.268 | 0.0031 | 0.0015 | 0.0029 | 0.0020 |
|  | 2015-16 | 100.729 | 0.0032 | 0.0016 | 0.0029 | 0.0021 |
|  | 2016-17 | 85.539 | 0.0032 | 0.0015 | 0.0029 | 0.0020 |
|  | 2017-18 | 85.853 | 0.0036 | 0.0016 | 0.0031 | 0.0020 |
|  | 2018-19 | 80.849 | 0.0032 | 0.0016 | 0.0030 | 0.0020 |
| Virginia | 2014-15 | 44.862 | 0.0032 | 0.0015 | 0.0029 | 0.0020 |
|  | 2015-16 | 107.501 | 0.0031 | 0.0016 | 0.0030 | 0.0022 |
|  | 2016-17 | 95.983 | 0.0032 | 0.0016 | 0.0031 | 0.0021 |
|  | 2017-18 | 84.540 | 0.0035 | 0.0016 | 0.0032 | 0.0020 |
|  | 2018-19 | 96.904 | 0.0032 | 0.0016 | 0.0031 | 0.0021 |

**Figure S2.**
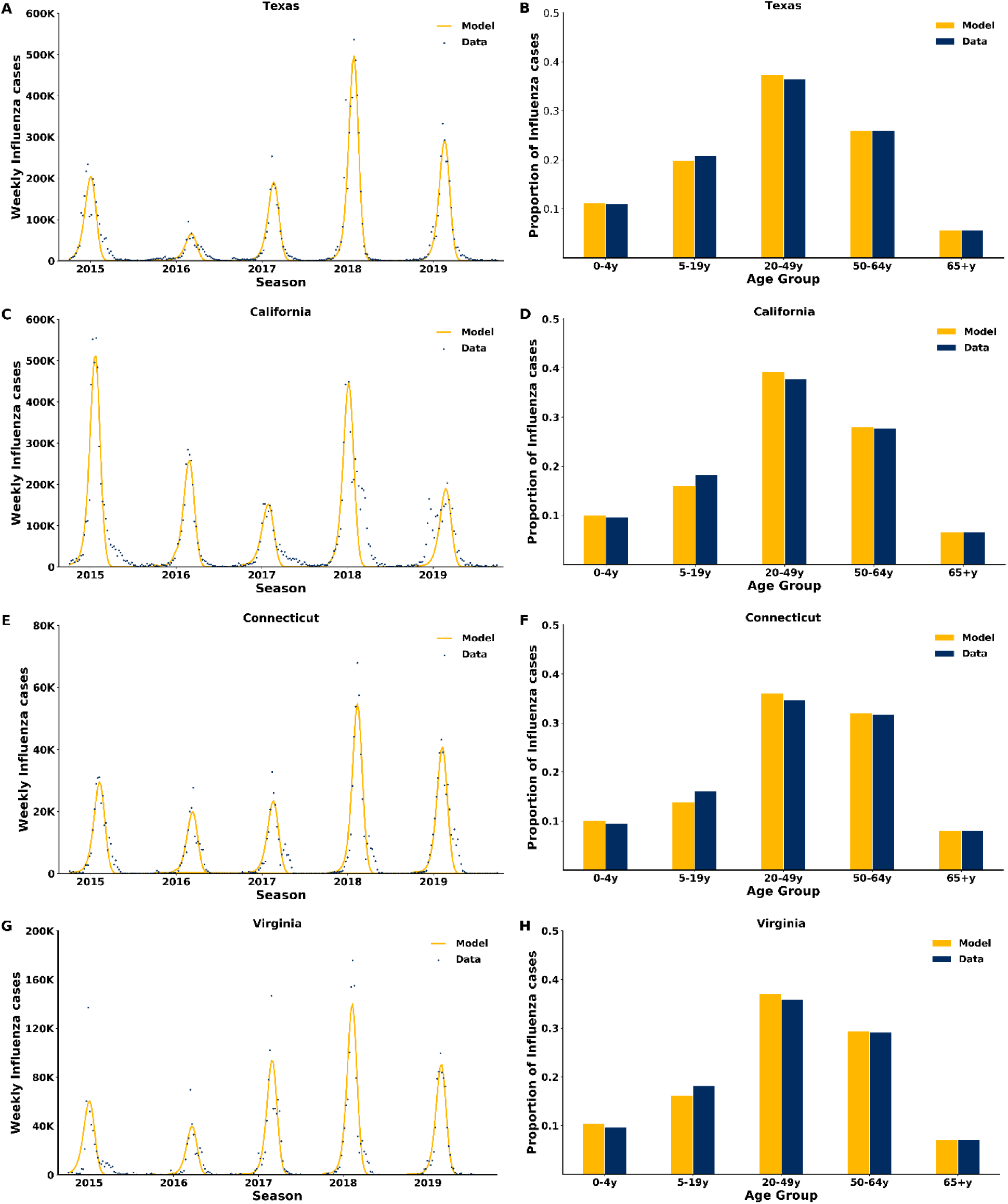
Model fit. Time series of recorded weekly influenza cases and model fit to Texas, California, Connecticut and Virginia (A, C, E & G). Data and model fit to the age distribution among influenza infections (B, D, F & H).

## 3. Model simulations

### Main text Figure 2

To evaluate the number of cases and hospitalizations averted by early treatment (within 48 hours since symptoms onset. For conservative purposes, we assumed treating on the second day) of infected high-risk individuals who are currently receiving treatment on the third and more than three days after symptoms onset (Main Text, Figure 2), we ran the model for each of the five seasons with an increased proportion of the infected high-risk treated on the second day since symptoms onset. For each season, we ran the model using the average state’s vaccination coverage. The addition to the proportion of the infected high-risk treated on the second day was done by shifting first the individuals who treated ineffectively (more than 72 hours since symptoms onset), followed by the individuals who treated on the third day.

The number of additional cases and hospitalizations averted was computed as the average over the model projections of 5 seasons compared to the baseline case. For the range, we did the same analysis described above but we used the vaccination coverage of the season with the highest (lower bond) and the lowest (higher bond) vaccination coverage in the seasons between (2013 - 2018) for each state.

### Main text Figure 3

We estimated the number of additional cases and hospitalizations averted due to increasing the treatment coverage of high-risk patients assuming that all receive treatment within 48 hours from symptoms onset. To assess the benefit in terms of cases and hospitalizations averted we ran the model with treatment coverage varying from 10%-30% for each season and state using the state’s average vaccination coverage. We also assumed that there are no infected high-risk individuals who getting treatment three days since symptoms onset. For each fixed portion we compared the mean model projections to the baseline scenario.

We also conducted a sensitivity analysis to examine the robustness of our results. In this analysis, we examined the effect of the vaccination coverage on the number of additional cases and hospitalizations averted by changing vaccination coverage for both model’s projections of the new treatment policy and the baseline treatment scenario. We increased and decrease the state’s mean vaccination coverage by 10%, 20%.

### Main text Figure 4

To estimate cases and hospitalizations averted per treatment for each age group we analyzed the scenario in which all the infected high-risk individuals in the examined age-group are seeking care and getting treatment within 48 hours since symptoms onset. For the rest of the age-groups we change the timeliness of initiating treatment to 48 hours since symptoms onset without changing the baseline treatment coverage. The number of additional cases and hospitalizations averted per treatment was computed by comparing the model’s projections of the new scenario with the baseline scenario, while considering the increase in the number of treated high-risk patients.

### Main text Figure 5

We conducted a two-way sensitivity analysis to investigate the joint impact of seasonal attack rate and vaccination coverage in each state on the number of cases averted per treatment. We ran the model with varying proportion of infected high-risk who seeks care and gets treated within 48 hours from symptoms onset from 10%-30% while increasing and decreasing the state’s average vaccination coverage by 10%-40%. We conducted the analysis for median, high, and low seasonal attack rate for each state. The high and low attack rate were informed by the year with the lowest and the year with the highest attack rate for each state between 2013-2018.

## 4. Additional results

**Figure S3.**
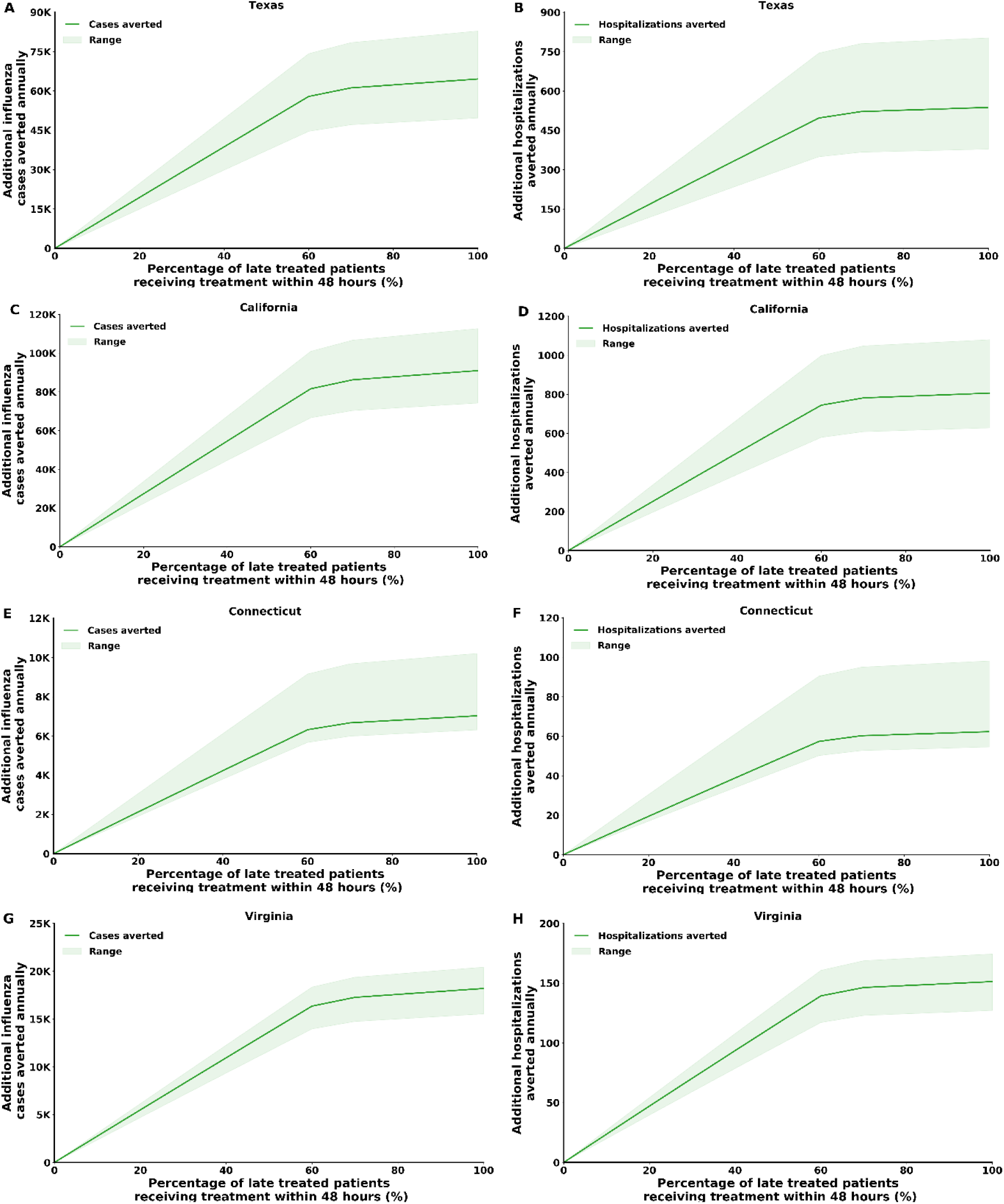
Model projection of in influenza cases and hospitalizations averted in Texas, California, Connecticut, and Virginia if treating within 48 hours from symptoms onset, 0-100% of the high-risk individuals who were treated after 48 hours. (A, C, E & G) Total number of cases averted. (B, D, F & H) Total number of hospitalizations averted.

**Figure S4.**
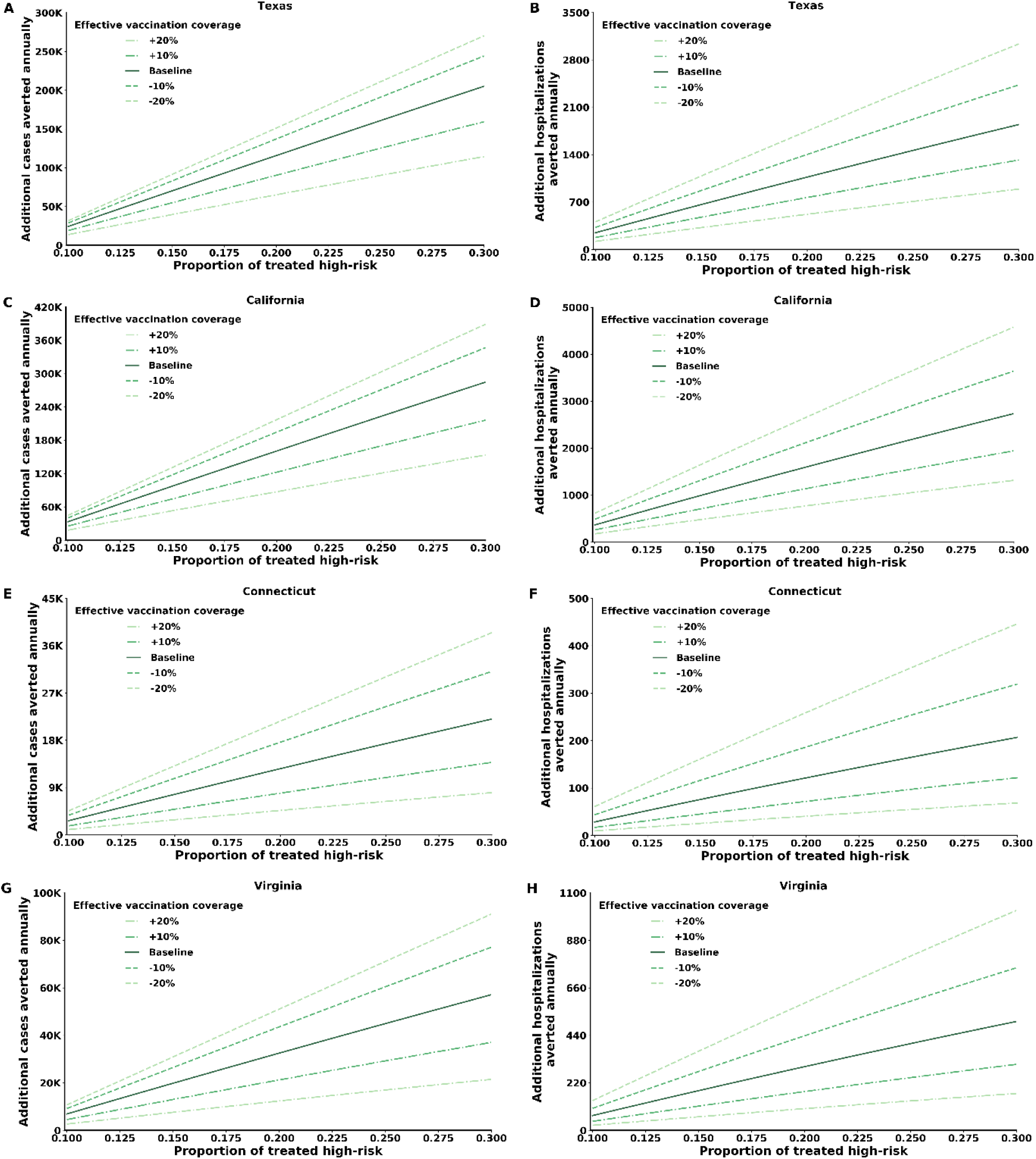
Model projections of influenza cases and hospitalizations averted in Texas, California, Connecticut, and Virginia by increasing the portion of treated high-risk individuals who seek care and get treated within 48 hours from symptoms onset. (A, C, E & G) Total number of cases averted. (B, D, F & H) Total number of hospitalizations averted.

**Figure S5.**
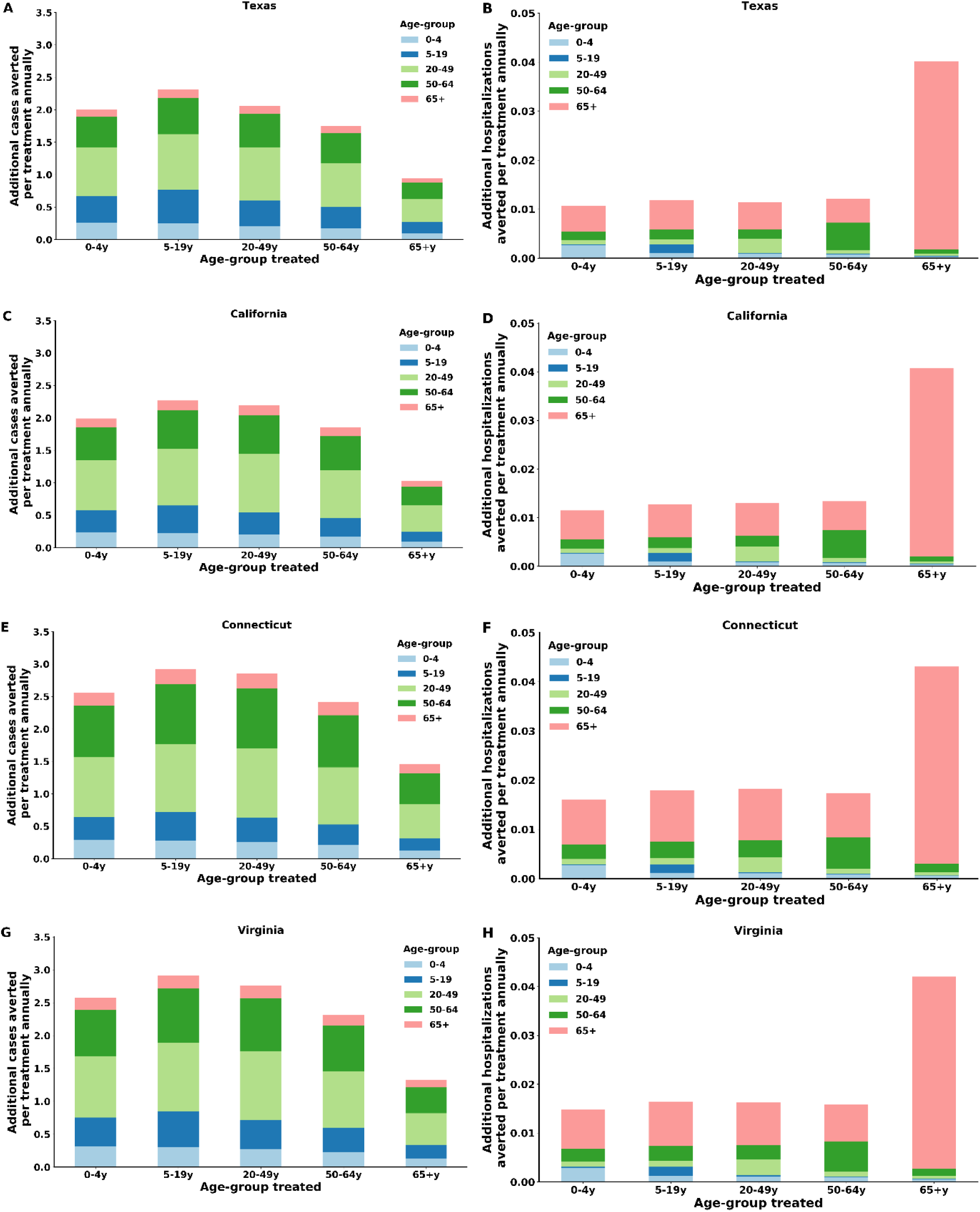
Model projections of influenza cases and hospitalizations averted per treatment, by treating each age-group 48 hours since symptoms onset in Texas, California, Connecticut and Virginia. (A, C, E & G) Number of cases averted per treatment for each age-group stratified by age-group. (B, D, F & H) Number of hospitalizations averted per treatment for each age-group stratified by age-group.

**Figure S6.**
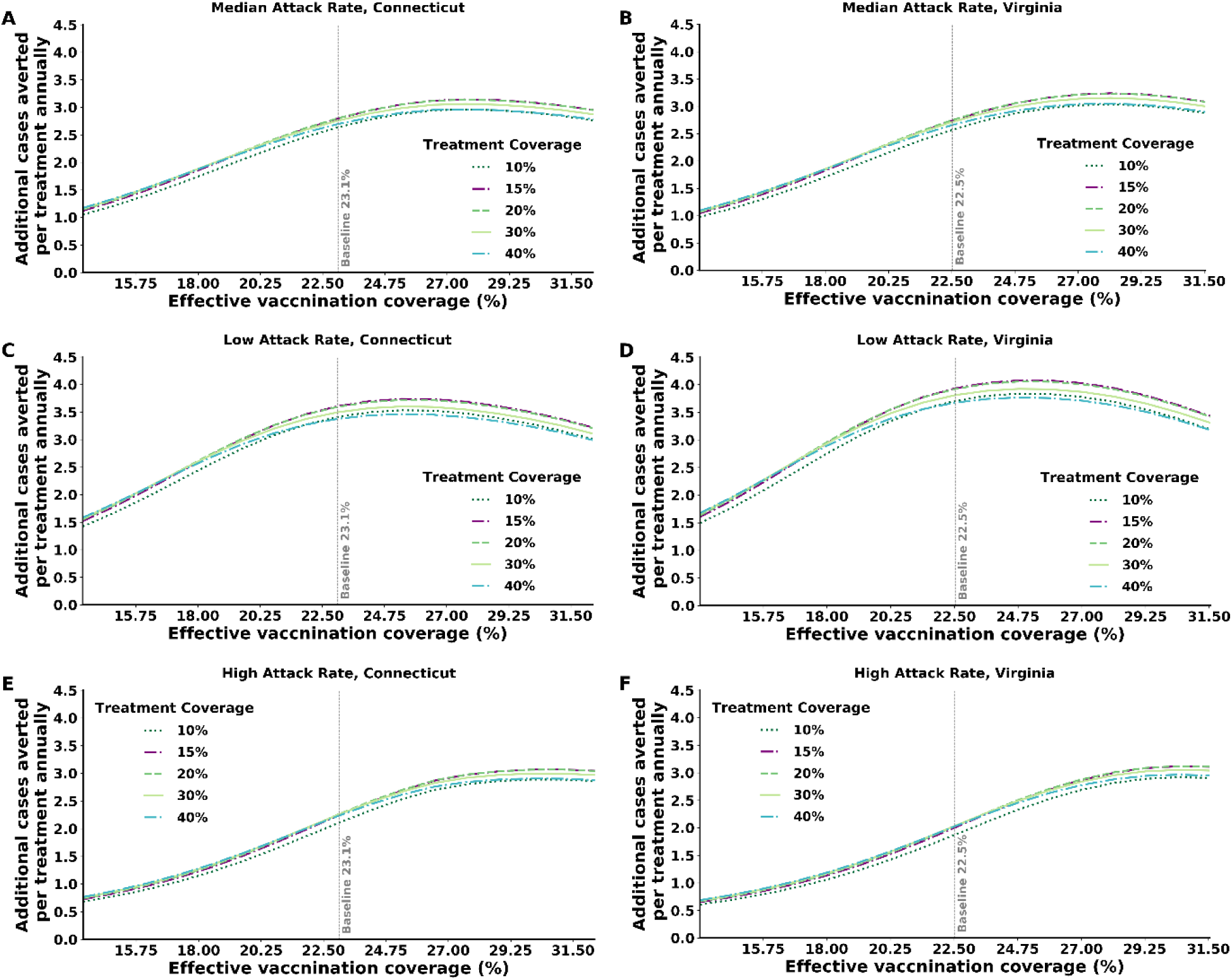
The mutual effect of attack rate and vaccination coverage, in Connecticut and Virginia, on the number of cases averted per treatment for each treatment coverage among high-risk individuals infected with influenza. Infected high-risk individuals seek care and get treatment within 48 hours from symptoms onset. (A, B) Median attack rate settings. (C, D) Low attack rate settings. (E, F) High attack rate settings.

